## Supplemental for "Wastewater surveillance in smaller college communities may aid future public health initiatives"

**PCR Performance QC**

The presence of PCR inhibitors in the sample matrix was tested by spiking 2 x 10^5^ copies of the IDT 2019-nCoV_N positive control plasmid into extracted wastewater in which no SARS-CoV-2 was detected and comparing the Cq to the same concentration of plasmid in the standard. There was no significant difference in Cq between the wastewater extract (24.87) and standard (24.76) suggesting no PCR inhibition.

The N1 primer set and the positive control plasmid generated a standard with an R^2^ value of 0.978 and 97.8% efficiency (slope −3.39, intercept 42.71). The N2 primer set and the positive control plasmid generated a standard with an R^2^ value of 0.991 and 95.9% efficiency (slope −3.46, intercept 43.549). The PMMoV primer set generated a standard with an R^2^ value of 0.996 and 88.7% efficiency (slope −3.65, intercept 42.15).

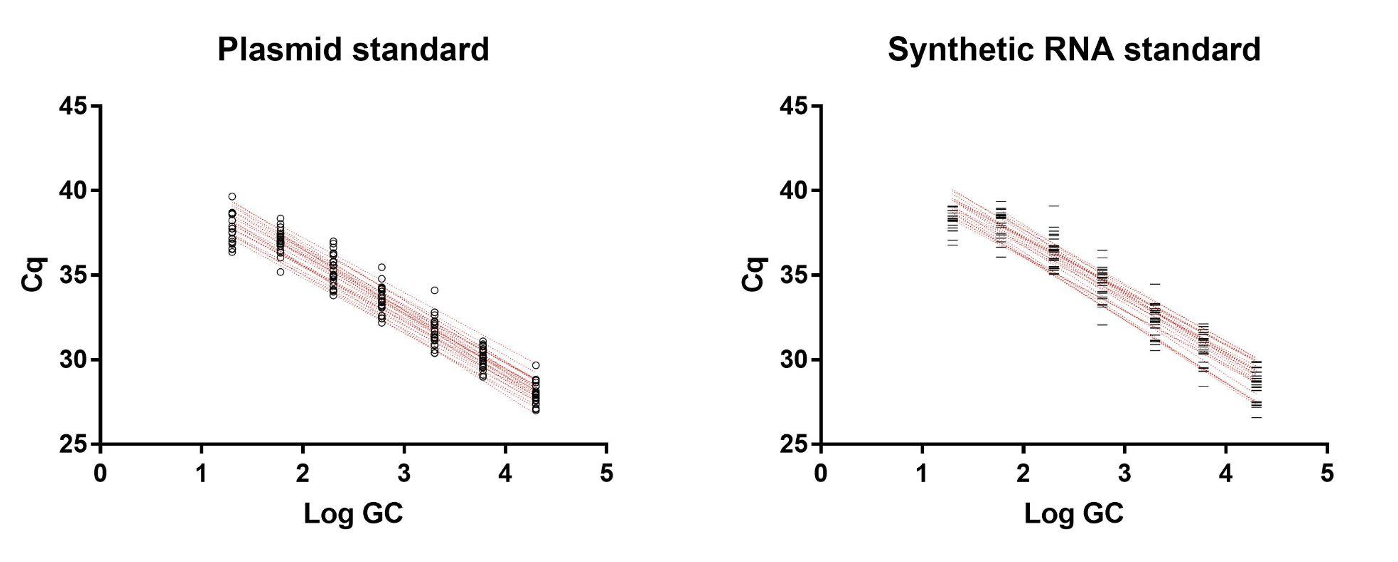
Plasmid standards have been reported to overestimate the viral load due to inefficient amplification of the supercoiled template during earlier cycles [23]. In our study, the synthetic RNA-derived standards were slightly less efficient (N1 93.7%, N2 90.1%) thus the positive control plasmid was used for quantification of gene copies.

***Supplemental Figure 1. Standard curves obtained from IDT 2019-nCoV_N positive control plasmid (left) and reverse-transcribed ATCC VR3276SD synthetic RNA as described in Methods.***

***Supplemental Table 1. Description of Collection and/or Detection Failure in Ruston, LA WBE***

| **Case Intervals** | **Wastewater Sample Dates** | **N1 Raw** | **N2 Raw** | **N1 normalized** | **N2 normalized** |
| --- | --- | --- | --- | --- | --- |
| **5/20-5/26** | **5/26/21** | 394000 | 12000 | 142.89 | 5.055 |
| **5/27-6/2** | **⊗** | **⊗** | **⊗** | **⊗** | **⊗** |
| **6/3-6/9** | **6/7/21** | 68000 | 18000 | 24.51 | 7.59 |
| **6/10-6/16** | **6/10/21** | 292000 | 180000 | 54.56 | 39.41 |
|  | **6/14/21** | **#** | 8000 | **#** | **#** |
| **6/17-6/23** | **6/17/21** | 400000 | 12000 | 39.74 | 1.41 |
|  | **6/21/21** | 124000 | 6000 | 360.7 | 19.98 |
| **6/24-6/30** | **⊗** | **⊗** | **⊗** | **⊗** | **⊗** |
| **7/1-7/7** | **⊗** | **⊗** | **⊗** | **⊗** | **⊗** |
| **7/8-7/14** | **7/12/21** | 56000 | 2000 | **×** | **×** |
|  | **7/14/21** | 130000 | 132000 | 31.97 | 38.27 |
| **7/15-7/21** | **7/19/21** | 550000 | 6000 | **×** | **×** |
|  | **7/21/21** | 254000 | 8000 | **×** | **×** |
| **7/22-7/28** | **7/26/21** | 350000 | 12000 | **×** | **×** |
|  | **7/28/21** | 1098000 | 30000 | 159.0 | 5.15 |
| **7/29-8/4** | **8/2/21** | 626000 | 8000 | **×** | **×** |
|  | **8/4/21** | 1488000 | 12000 | 412.32 | 3.94 |
| **8/5-8/11** | **8/9/21** | 392000 | 500000 | 53.81 | 80.92 |
| **8/12-8/18** | **8/12/21** | 18000 | 78000 | 1.98 | 10.78 |
|  | **8/17/21** | 1886000 | 24000 | 6166.73 | 95.09 |
| **8/19-8/25** | **8/19/21** | 8000 | 126000 | 2.09 | 34.83 |
|  | **8/23/21** | 300000 | 22000 | 949.06 | 84.77 |
| **8/26-9/1** | **8/30/21** | 60000 | 2208000 | 7.01 | 304.57 |
| **9/2-9/8** | **⊗** | **⊗** | **⊗** | **⊗** | **⊗** |
| **9/9-9/15** | **9/13/21** | 242000 | 18000 | 671.24 | 58.7 |
|  | **9/15/21** | 220000 | 16000 | 37.38 | 3.05 |
| **9/16-9/22** | **⊗** | **⊗** | **⊗** | **⊗** | **⊗** |
| **9/23-9/29** | **9/27/21** | 274000 | 62000 | 54.07 | 14.35 |
| **9/30-10/6** | **9/30/21** | 194000 | 4000 | 739.62 | 13.77 |
|  | **10/4/21** | 410000 | 6000 | 2218.4 | 41.76 |
| **10/7-10/13** | **⊗** | **⊗** | **⊗** | **⊗** | **⊗** |
| **10/14-10/20** | **⊗** | **⊗** | **⊗** | **⊗** | **⊗** |
| **10/21-10/27** | **10/21/21** | 8000 | 620 | 24.1 | 2.34 |
| **10/28-11/3** | **10/27/21** | 12000 | 2000 | 40.21 | 10.21 |
| **11/4-11/10** | **11/10/21** | 38000 | 54000 | 4.47 | 7.7 |
| **11/11-11/17** | **11/16/21** | 52000 | 82000 | 6.41 | 11.71 |
| **11/18-11/24** | **⊗** | **⊗** | **⊗** | **⊗** | **⊗** |
| **11/25-12/1** | **⊗** | **⊗** | **⊗** | **⊗** | **⊗** |
| **12/2-12/8** | **⊗** | **⊗** | **⊗** | **⊗** | **⊗** |
| **12/9-12/15** | **⊗** | **⊗** | **⊗** | **⊗** | **⊗** |
| **12/16-12/22** | **12/21/21** | 14000 | 16000 | 84.3 | 117.58 |
| **12/23-12/29** | **12/28/21** | 12000 | 298000 | 1.0 | 28.99 |
| **12/30-1/5** | **⊗** | **⊗** | **⊗** | **⊗** | **⊗** |
| **1/6-1/12** | **1/12/22** | 6000 | 12000 | 16.06 | 28.99 |
| **1/13-1/19** | **1/19/22** | 2000 | 6000 | 156.13 | 772.67 |
| **1/20-1/26** | **1/26/22** | 24000 | 70000 | 175.57 | 604.07 |
| **1/27-2/2** | **2/2/22** | 20000 | 56000 | 57.34 | 198.26 |
| **2/3-2/9** | **2/9/22** | 16000 | 12000 | 88.32 | 74.28 |
| **2/10-2/16** | **2/16/22** | 10000 | 720 | 556.95 | 60.0 |
| **2/17-2/23** | **⊗** | **⊗** | **⊗** | **⊗** | **⊗** |
| **2/24-3/2** | **3/2/22** | 14000 | 6000 | 4.43 | 2.18 |
| **3/3-3/9** | **3/9/22** | 22000 | 8000 | 8.35 | 3.75 |
| **3/10-3/16** | **⊗** | **⊗** | **⊗** | **⊗** | **⊗** |
| **3/17-3/23** | **3/22/22** | 432000 | 4000 | 968.19 | 8.34 |
| **3/24-3/30** | **3/30/22** | 1322000 | 302000 | 487.13 | 130.68 |
| **3/31-4/6** | **4/6/22** | 694000 | 26000 | 65.15 | 2.93 |
| **4/7-4/13** | **4/13/22** | 1300000 | 170000 | 269.34 | 41.51 |
| **4/14-4/20** | **⊗** | **⊗** | **⊗** | **⊗** | **⊗** |
| **4/21-4/27** | **4/27/22** | 22000 | 6000 | 20.89 | 5.89 |
| **4/28-5/4** | **5/4/22** | 4000 | 4000 | 1.14 | 1.0 |
| **⊗** Denotes a week without wastewater sample collection from WWTP | | | | | |
| **#** Denotes where SARS-CoV-2 data could not be detected due to insufficient WW volume | | | | | |
| * Denotes where PMMoV data could not be collected due to insufficient WW volume | | | | | |
